# Characterizing key attributes of the epidemiology of COVID-19 in China: Model-based estimations

**DOI:** 10.1101/2020.04.08.20058214

**Authors:** Houssein H. Ayoub, Hiam Chemaitelly, Ghina R Mumtaz, Shaheen Seedat, Susanne F. Awad, Monia Makhoul, Laith J. Abu-Raddad

**Author notes:** Reprints or correspondence: Dr. Houssein H. Ayoub, Department of Mathematics, Statistics, and Physics, Qatar University, P.O. Box 2713, Doha, Qatar., Professor Laith J. Abu-Raddad, Infectious Disease Epidemiology Group, Weill Cornell Medicine - Qatar, Qatar Foundation - Education City, P.O. Box 24144, Doha, Qatar.

## Abstract

**Background:** A novel coronavirus strain, severe acute respiratory syndrome coronavirus 2 (SARS-CoV-2), emerged in China in late 2019. The resulting disease, Coronavirus Disease 2019 (COVID-2019), soon became a pandemic. This study aims to characterize key attributes of the epidemiology of this infection in China.

**Methods:** An age-stratified mathematical model was constructed to describe the transmission dynamics and estimate the age-specific differences in the biological susceptibility to the infection, age-assortativeness in transmission mixing, case fatality rate (CFR), and transition in rate of infectious contacts (and reproduction number *R*_0_) following introduction of mass interventions.

**Results:** The model estimated the infectious contact rate in early epidemic at 0.59 contacts per day (95% uncertainty interval (UI)=0.48-0.71). Relative to those 60-69 years of age, susceptibility to the infection was only 0.06 in those ≤19 years, 0.34 in 20-29 years, 0.57 in 30-39 years, 0.69 in 40-49 years, 0.79 in 50-59 years, 0.94 in 70-79 years, and 0.88 in ≥80 years. The assortativeness in transmission mixing by age was very limited at 0.004 (95% UI=0.002-0.008). Final CFR was 5.1% (95% UI=4.8-5.4%). *R*_0_ rapidly declined from 2.1 (95% UI=1.8-2.4) to 0.06 (95% UI=0.05-0.07) following onset of interventions.

**Conclusion:** Age appears to be a principal factor in explaining the patterns of COVID-19 transmission dynamics in China. The biological susceptibility to the infection seems limited among children, intermediate among young to mid-age adults, but high among those >50 years of age. There was no evidence for differential contact mixing by age, consistent with most transmission occurring in households rather than in schools or workplaces.

## Introduction

An outbreak of a novel coronavirus strain, severe acute respiratory syndrome coronavirus 2 (SARS-CoV-2), was identified in Wuhan, Hubei province, China, in late December 2019 [1, 2]. The outbreak started with identification of four cases of severe pneumonia of unknown etiology, but with symptoms similar to those of Severe Acute Respiratory Syndrome (SARS) and the Middle East Respiratory Syndrome (MERS) [1, 3]. Initial cases were linked to exposure at the Huanan Seafood Market, but subsequent infections resulted from rapid community transmission [1-3]. Within about two months, over 80,000 cases and 3,000 deaths occurred across China [2, 4], amid extreme preventive measures and outstanding healthcare mobilization [2, 3]. The resulting disease was named Coronavirus Disease 2019 (COVID-2019) by the World Health Organization (WHO) [5], and has been declared a pandemic [6] after affecting tens of countries and territories [4]. For simplicity, we will thereafter refer to this virus as “COVID-19”, though it is the name of the disease form, given its prevalent current use in the public sphere and to avoid confusion between SARS-CoV-2 and SARS-CoV.

The aims of this study are to investigate and characterize key attributes of the epidemiology of COVID-19 in China including 1) age-specific differences in the *biological* susceptibility to infection, 2) age-assortativeness in infection transmission, and 3) transition in the rate of infectious contacts (and reproduction number) following the introduction of mass interventions.

## Materials and Methods

### Mathematical Model

A deterministic compartmental mathematical model was constructed to describe COVID-19 transmission dynamics in a given population, and was applied here to the population of China (Figure S1 of Supplementary Information (SI)). The model was designed based on current understanding of the infection’s natural history and epidemiology. The model consists of a set of coupled nonlinear differential equations that stratify the population into compartments according to age group, infection status, infection stage, and disease stage. Analyses were performed in MATLAB R2019a [7].

The model stratified the population into nine age groups, each representing a ten-year age band except for the last category (0-9, 10-19, …, ≥80 years). Susceptible individuals in each age group are at risk of being exposed to the infection at varying hazard rates, which are age-and time-dependent, to capture the variability in the risk of exposure and the impact of public health interventions. Following a latency period, infected individuals are stratified to develop mild infection followed by recovery, or severe infection followed by severe disease then recovery, or critical infection followed by critical disease and either recovery or disease mortality.

The model parameterized the variation in the rate of infectious contacts through a Woods-Saxon function [8-11] to characterize the transition after China’s robust public health response in terms of its scale or strength, smoothness or abruptness, duration, and the turning point. The model also incorporated an age mixing matrix that allows a range of contact mixing between individuals varying from fully assortative (mixing only with individuals in the same age group) to fully proportionate (mixing with individuals with no preferential bias for a specific age group). The latter was incorporated to capture any assortativeness in infection transmission mixing by age. Further details on model structure can be found in Section 1 of SI.

### Model Parameterization

The model was parameterized using current data on COVID-19 natural history and epidemiology. The duration of latent infection was set at 3.69 days based on an existing estimate [12] and based on a median incubation period across confirmed cases of 5.1 days [1], adjusted for the observed viral load among infected persons following exposure [13] and reported infection transmission prior to onset of symptoms [14]. The age-stratified proportions of infected individuals that will eventually progress to develop mild, severe, or critical infections were based on the observed distribution of cases across these infection stages in China [3, 15, 16]. The duration of infectiousness was assumed to last for 3.48 days based on an existing estimate [12] and based on the observed time to recovery in persons with mild infection [3, 12] and the observed viral load among infected persons [13, 14].

Individuals with severe (or critical) infections develop severe (or critical) disease over a period of 28 days prior to recovery, as informed by the observed duration from onset of severe (or critical) disease to recovery [3]. Individuals with critical disease had the additional risk of disease mortality [17]. The disease mortality rate was fitted factoring the observed crude case fatality rate in each age group in China as of February 11, 2020 [2, 17].

The population size, demographic structure (age distribution), and life expectancy of the population of China, as of 2020, were obtained from the United Nations World Population Prospects database [18].

Further details on model parameters, values, and justifications can be found in Tables S1 and S2 and Section 2 of SI.

### Model Fitting

The model was fitted to the following sources of data: 1) time series of diagnosed COVID-19 cases and of the cumulative number of diagnosed COVID-19 cases and of recovered individuals [3], 2) time series of reported COVID-19 deaths and of the cumulative number of COVID-19 deaths [3], and 3) relative attack rate by age, that is the proportion of the population that has already been infected by February 11, 2020 stratified by age [17]. China’s reported cases and deaths were adjusted to reflect the change in coronavirus case definition to include, in addition to the laboratory-confirmed cases, those who are clinically-diagnosed [19, 20] (Section 2 of SI).

Model fitting was used to estimate the infectious contact rate, age-stratified susceptibility to the infection, degree of assortativeness in the age group mixing, overall attack rate, overall disease mortality rate, time delay between onset of actual infection and case notification, and between actual death and reported death, and transition in the basic reproduction number *R*_0_. A nonlinear least-square data fitting method, based on the Nelder-Mead simplex algorithm, was used to minimize the sum of squares between data points and model predictions [21].

### Uncertainty Analyses

A multivariable uncertainty analysis was conducted to determine the range of uncertainty around model predictions. Five-hundred simulation runs were performed, applying at each run, Latin Hypercube sampling from a multidimensional distribution of the model parameters, where parameter values are selected from ranges specified by assuming ±30% uncertainty around parameters’ point estimates. The model was then refitted to the input data, and the resulting distributions of estimates, across all 500 runs, were used to calculate the model predictions’ means and 95% uncertainty intervals (UIs).

## Results

The model fitted the different COVID-19 empirical data such as time-series of diagnosed cases (Figure 1A), time-series of reported deaths (Figure 1B), and age-stratified attack rate as of February 11, 2020 (Figure 1C). The model estimated the epidemic emergence at ∼49 days (95% UI: 48-50) prior to January 17, 2020, that is towards the end of November, 2019. At the beginning of the epidemic, the predicted infectious contact rate was 0.59 contacts per day (95% UI: 0.48-0.71; Figure S2D of SI). The predicted (average) time delay was 5.4 days (95% UI: 5.2-5.6) between onset of actual infection and reported infection, and 1.6 days (95% UI: 1.5-1.7) between actual death and reported death.

**Figure 1.**
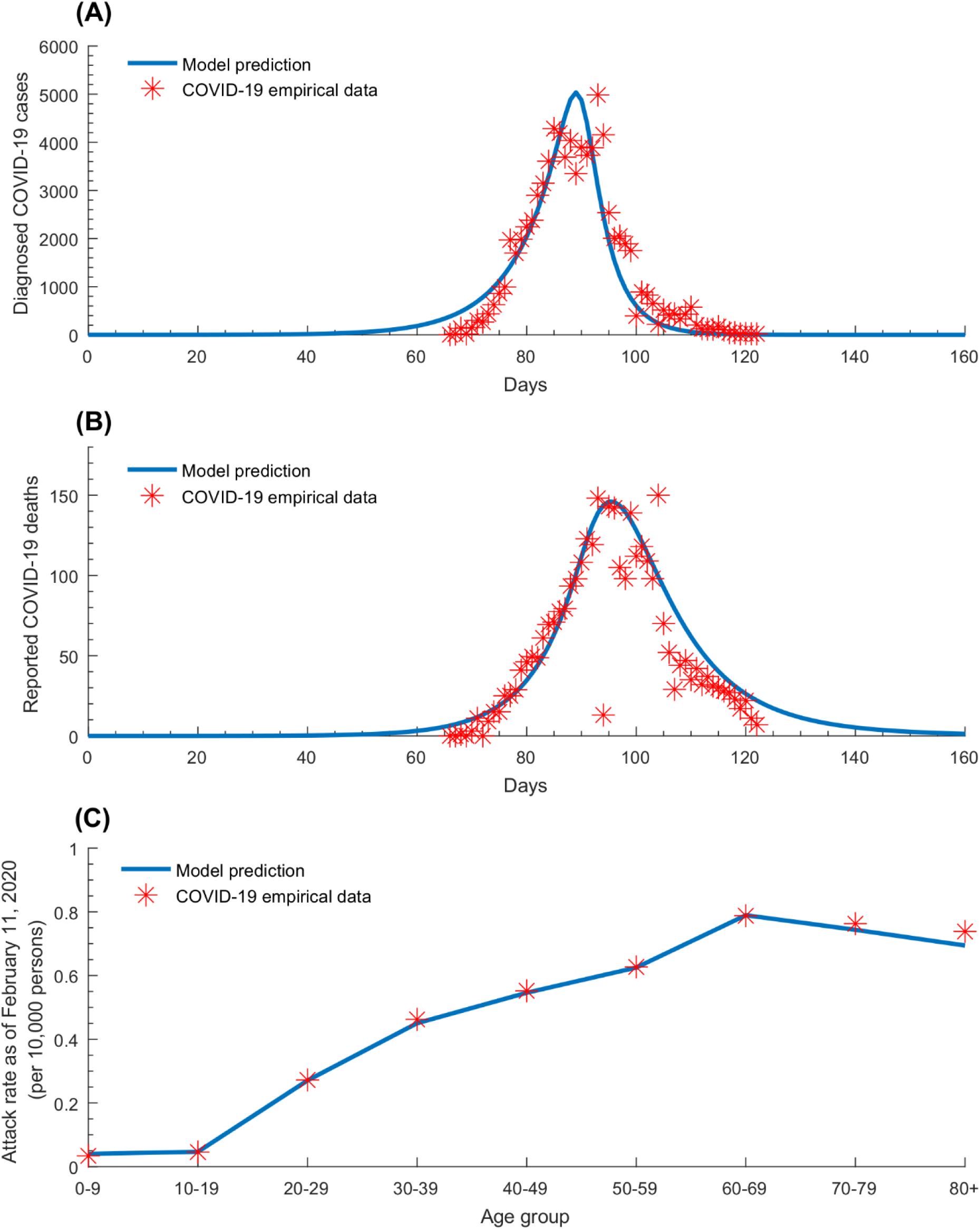
Model fitting of COVID-19 empirical data. Model fits to (A) the time-series of diagnosed cases, (B) the time-series of reported deaths, and (C) the age-stratified attack rate as of February 11, 2020.

Figure 2 shows the predicted time evolution of COVID-19 *crude* case fatality rate (CFR). In the early phase of the epidemic, the crude CFR increased rapidly following the rise in incidence, but plateaued shortly after (towards the end of the first month) and remained so till incidence reached its peak. When incidence started declining (∼90 days), the crude CFR grew rapidly, eventually saturating at ∼150 days. The *true* (that is final) CFR in the total population, estimated through the 500 simulation runs, was 5.1% (95% UI=4.8-5.4%; Figure S2A of SI).

**Figure 2.**
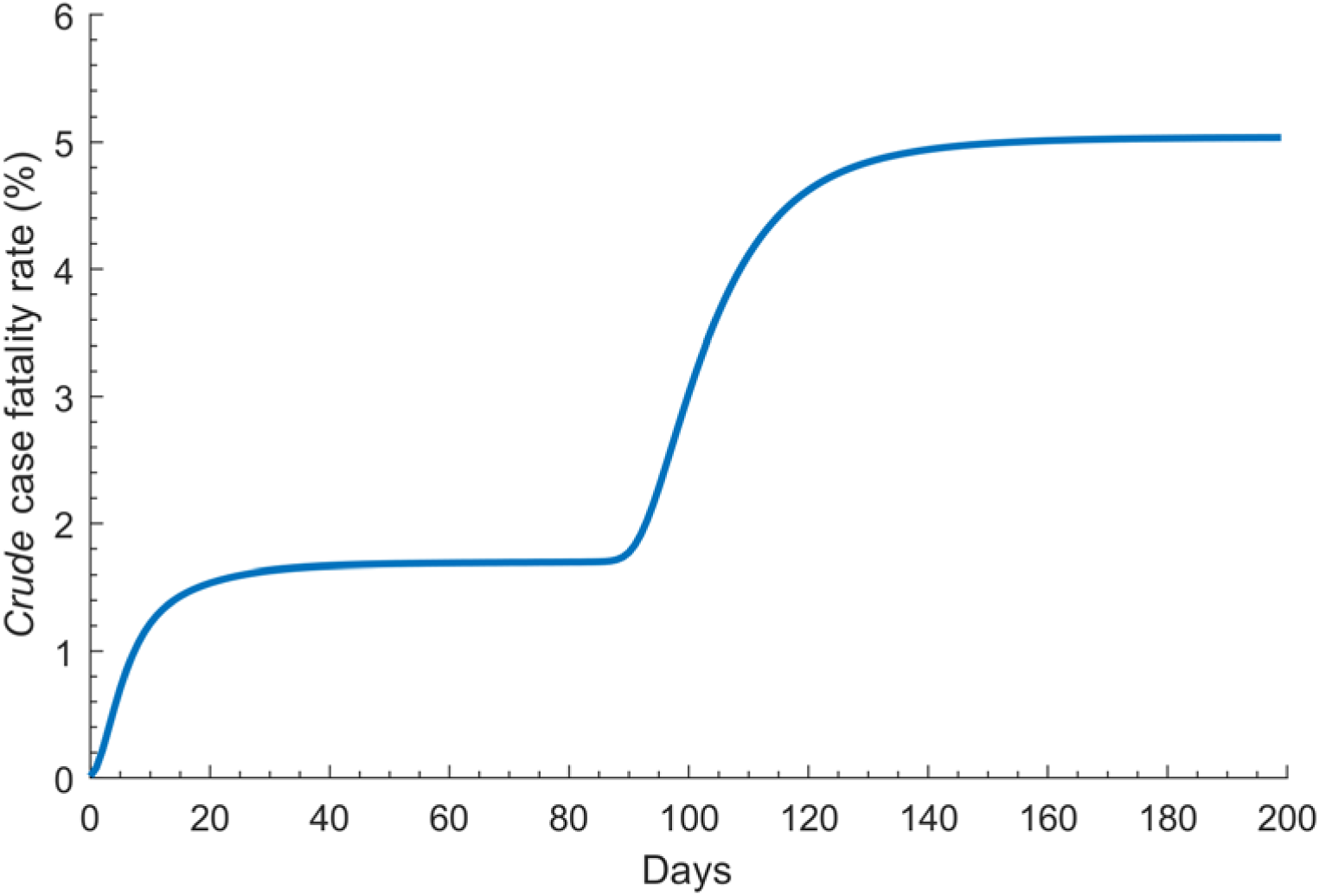
Model predictions for the time evolution of the COVID-19 *crude* case fatality rate.

Figure 3 features the estimated age-stratified susceptibility profile to COVID-19 infection. Susceptibility was lowest in individuals 0-9 years of age and highest in the 60-69 years age group. Relative to those 60-69 years of age, susceptibility to the infection was only 0.05 in those 0-9 years of age and 0.06 in those 10-19 years of age, but 0.34 in those 20-29 years of age, 0.57 in those 30-39 years of age, 0.69 in those 40-49 years of age, 0.79 in those 50-59 years of age, 0.94 in those 70-79 years of age, and 0.88 in those ≥80 years of age. The uncertainty analysis affirmed these results with narrow uncertainty intervals (Figure S2B of SI).

**Figure 3.**
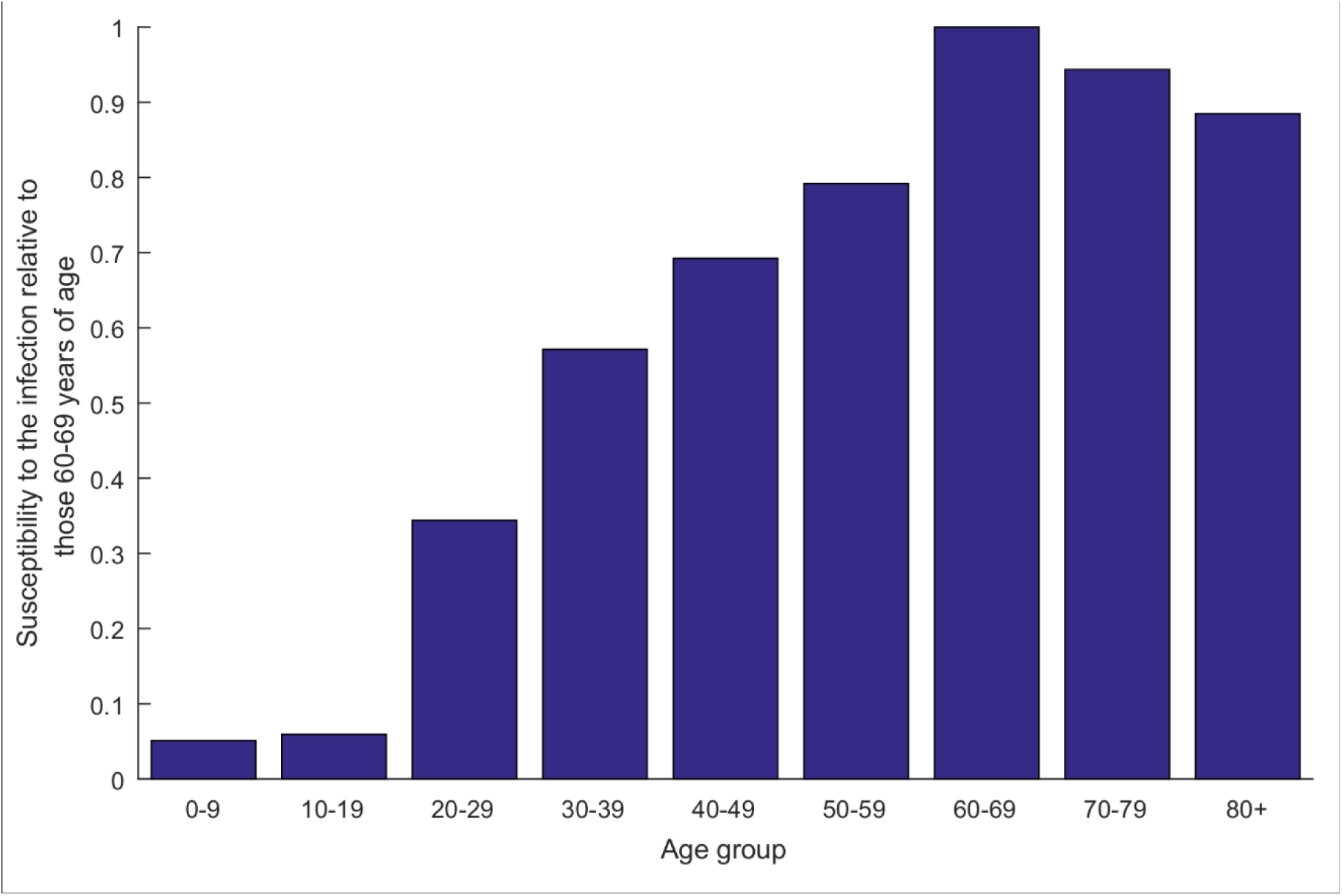
Model predictions for the age-stratified susceptibility profile to COVID-19 infection, *relative* to those 60-69 years of age.

Figure 4 illustrates the predicted degree of assortativeness in the age group mixing across the 500 simulation runs, which was estimated at 0.004 (95% UI=0.002-0.008)—there was virtually no assortativeness in infection transmission mixing by age.

**Figure 4.**
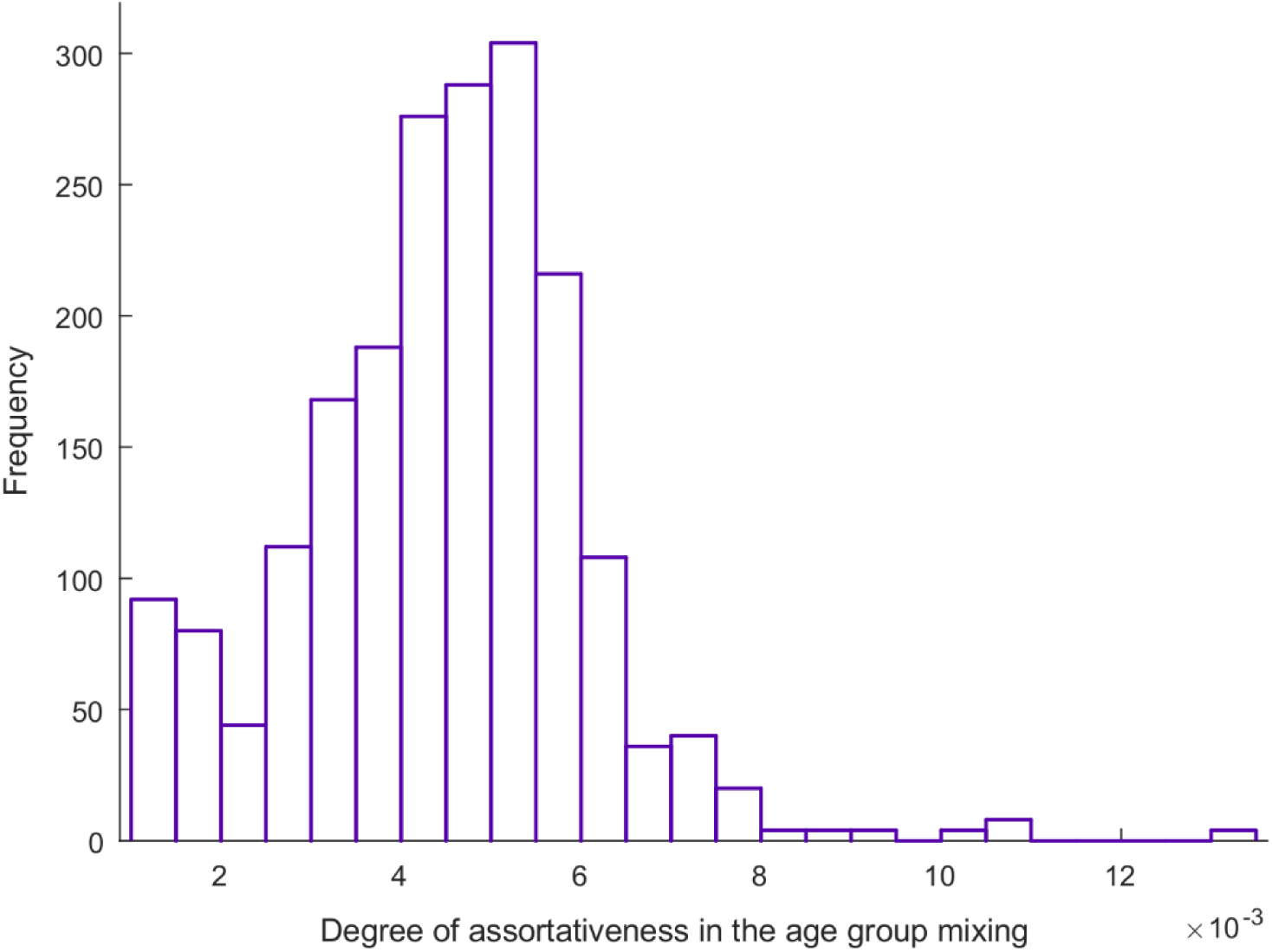
Model predictions for the degree of assortativeness in infection transmission mixing by age across the 500 uncertainty analysis simulation runs.

Figure 5 and Figure S2C of SI show the time evolution of *R*_0_, and its predicted mean and 95% UI across the 500 uncertainty runs, respectively. In the early phase of the epidemic, *R*_0_ was estimated at 2.1 (95% UI: 1.8-2.4), but rapidly declined to 0.06 (95% UI: 0.05-0.07) following the onset of interventions. The sharp transition duration for *R*_0_ was estimated at 11.5 days (95% UI: 9.5-13.0).

**Figure 5.**
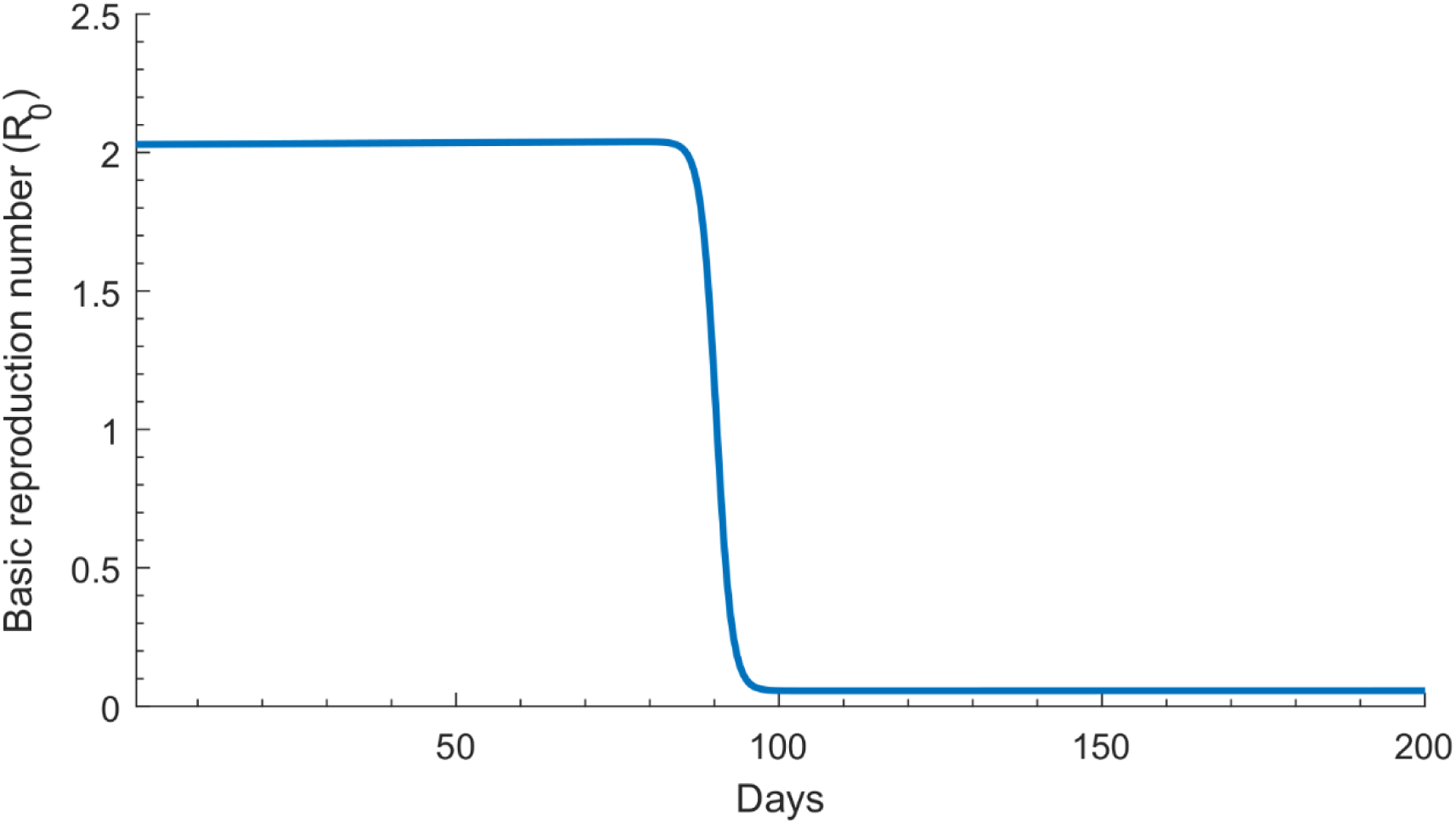
Model predictions for the time evolution of the basic reproduction number *R*_0_ before and after onset of the interventions in China.

## Discussion

Several key attributes of the epidemiology of COVID-19 have been investigated and estimated. A finding is that the *biological* susceptibility to the infection appears to vary by age (Figure 3). Susceptibility to COVID-19 was substantially higher among those >50 years of age compared to those in the younger age groups. For instance, compared to those 60-69 years of age, those ≤19 years of age, 20-29 years of age, and 30-39 years of age were, respectively, 94%, 68%, and 43% less susceptible to being infected. Notably, this age-dependence in the susceptibility to the infection could not be explained by differences in mixing between age groups, as the results indicated limited assortativeness in infection transmission mixing by age (Figure 4).

These findings support an important role for age in the epidemiology of this infection and affirm other studies suggesting lower susceptibility to the infection at younger age [22, 23].

Remarkably, the observed attack rate pattern for COVID-19 by age (Figure 1C) is the inverse (or better complement) of the age-specific cumulative incidence pattern of the 2009 influenza A (H1N1) pandemic (H1N1pdm) infection (Figure S3 of SI) [24]. This may suggest that a potential protective factor against COVID-19 acquisition (or rapid clearance) at young age could be prior recent exposure (cross immunity) to other viruses, particularly other cold coronaviruses [25]. An alternative hypothesis has suggested immune imprinting to a similar virus among adults [22]. This being said, the underlying immunological and/or epidemiological factors driving this age effect remain to be investigated with several alternative mechanisms potentially explaining this pattern. For instance, children and young adults may have subclinical infection with low viral load and rapid clearance with limited transmission potential to others. Of note that the contact tracing data from China suggest that children do not appear to play a significant role in the transmission [3, 26, 27].

The study predicted that the CFR will reach about 5% by the end of the outbreak in China (Figure 2), that is by the time all currently *active* cases would have been resolved, either through recovery or death. Our results indicated that the *crude* CFR observed in the first three months of the outbreak underestimated the *true* CFR by about 50%. This is because most infections were still recent infections and have not yet progressed to critical disease or death—death is a *late* outcome that was estimated to occur 2-8 weeks after onset of symptoms [3].

Another finding of this study is the limited assortativeness in infection transmission mixing by age (Figure 4), that is equal mixing between the different age groups. This finding is possibly explained by the fact that most transmissions occurred in the context of households rather than of schools or workplaces, as supported by existing evidence [3]. Indeed, analysis of 344 clusters in Guangdong and Sichuan provinces indicated that 78-85% of clusters pertained to families [3]. This finding is also supported by analysis of the contacts stratified by age in China [23]. It is, however, important to highlight that while this finding may apply to the Hubei outbreak (given the lockdown measures that may have limited all other transmission pathways), it may not necessarily be generalizable to other settings.

The present study affirmed the impact and success of the drastic preventive measures in curtailing infection transmission. *R*_0_ was sharply reduced by 97% over a short duration (Figure 5). At the beginning of the epidemic, on average, each infected person had 0.6 *infectious* contacts per day, that is each person passed the infection to 0.6 persons per day (Figure S2D of SI). The rate of infectious contacts can be expressed roughly as *c* × *p*, where *c* is the number of “social” contacts conducive to COVID-19 transmission per day and *p* is the transmission probability of the virus in a single contact. While *p* is unknown, it is possibly in the order of 1-5%, as suggested by contact tracing data from China—1-5% of contacts were subsequently laboratory-confirmed as COVID-19 cases [3]. This implies that, on average, each infected person had somewhere between 10-60 contacts per day at the beginning of the epidemic, but very few contacts after the lockdown. The latter further affirms the role of the lockdown in severely cutting the contact rate, making sustainable infection transmission very difficult.

This study has limitations. Model projections are contingent on the quality and representativeness of the input data. For instance, we assumed infection levels to be as officially documented, but evidence suggested that many infections may have been undocumented, particularly in the early phase of the outbreak [12]. The natural history of this infection is not yet firmly established, and the case management protocols have evolved over time [19, 20].

Mortality data seem to suggest that the standard of care improved over time especially in recent weeks when the healthcare sector was no longer overwhelmed with a large case load. We assumed a time-independent risk of disease mortality, and therefore we may have overestimated the true (final) CFR. We used a deterministic compartmental model, but this type of model may not be representative of the stochastic transmission dynamics when the number of infections is small, such as in the very early phase of the epidemic, thereby adding uncertainty to our estimate for the day of outbreak emergence. Despite these limitations, our parsimonious model, tailored to the nature of available data, was able to reproduce the COVID-19 epidemic as observed in China, and provided insights about infection transmission and disease progression in the population.

## Conclusion

Age appears to be a principal factor in explaining the patterns of COVID-19 transmission dynamics in China. The biological susceptibility to the infection seems limited among children, intermediate among young adults and those mid-age, but high among those >50 years of age. There was no evidence for considerable differential contact mixing by age, consistent with most transmission occurring in households rather than in schools or workplaces. Further mathematical modeling research is needed to understand the COVID-19 epidemics in other countries, and to draw further inferences about the global epidemiology of this infection.

## Data Availability

All data are within the paper and its Supplementary Information.

## Authors’ contributions

HHA constructed, coded, and parameterized the mathematical model and conducted the analyses. HC supported model parametrization and wrote the first draft of the paper. LJA conceived and led the design of the study, construct and parameterization of the mathematical model, and drafting of the article. All authors contributed to discussion and interpretation of the results and to the writing of the manuscript. All authors have read and approved the final manuscript.

## Availability of data and materials

All data are within the paper and its Supplementary Information.

## Ethics approval and consent to participate

Not applicable

## 1. Mathematical model

We developed a deterministic compartmental mathematical model to describe the transmission dynamics and disease progression of the severe acute respiratory syndrome coronavirus 2 (SARS-CoV-2) in a population (Figure S1). For simplicity, we will thereafter refer to this virus as COVID-19, given prevalent current use. This model stratifies the population into compartments according to age group (0-9, 10-19, 20-29,…, ≥80 years), and infection status (uninfected, infected), infection stage (mild, severe, critical), and disease stage (severe, critical).

**Figure S1.**
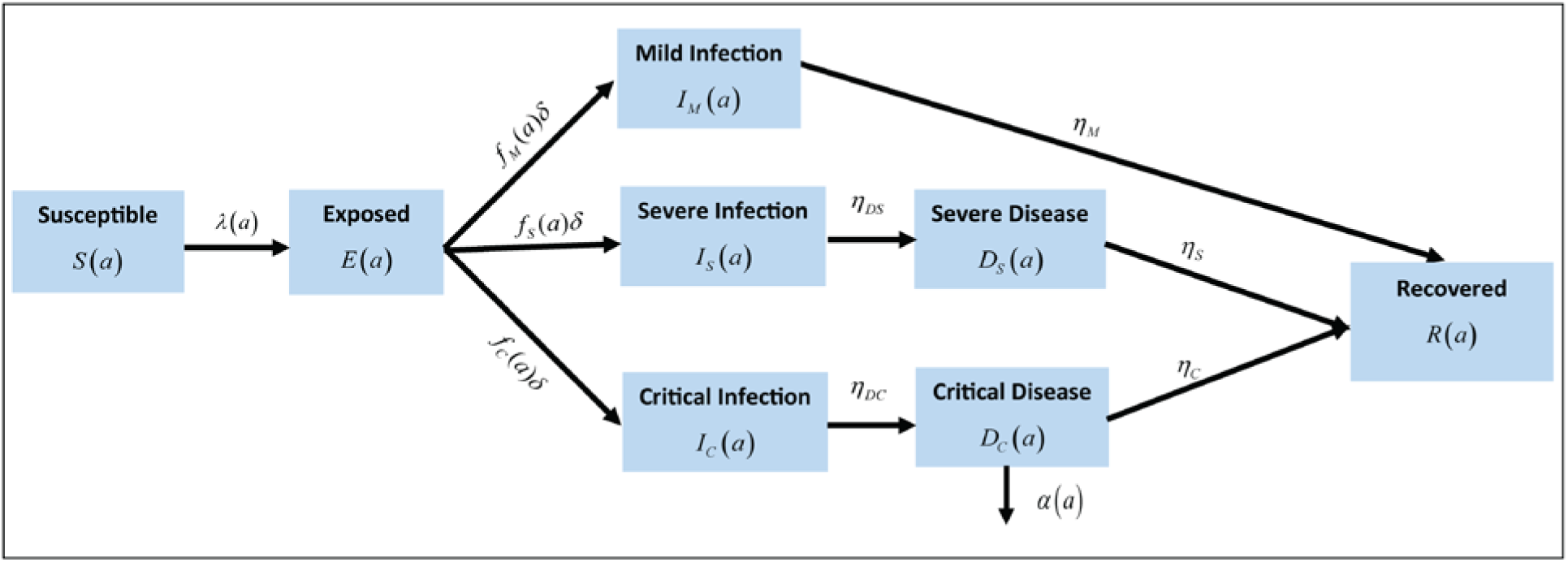
Schematic diagram describing the basic structure of the COVID-19 model.

### 1.1. Model equations and description

The model was expressed in terms of a system of coupled nonlinear differential equations for each age group. The index *a* denotes the different age cohorts (*a* = 1, 2,…, 9) in the population, with each age group representing a ten-year age band apart from the last one (those ≥80 years of age). The population size and demographic structure were set by China’s demography as provided by the United Nations World Population Prospects database [1]. However, the population size and distribution across age groups were fixed at 2020 levels, to disentangle the epidemiologic and demographic effects—a valid assumption given that the current time scale of the epidemic is only few months. All COVID-19 mortality was assumed to occur in individuals that are in the critical disease stage, as informed by the China outbreak data [2].

The following set of equations was used to describe the epidemic dynamics in the first age group:

<u>Population aged 0-9 years:</u>

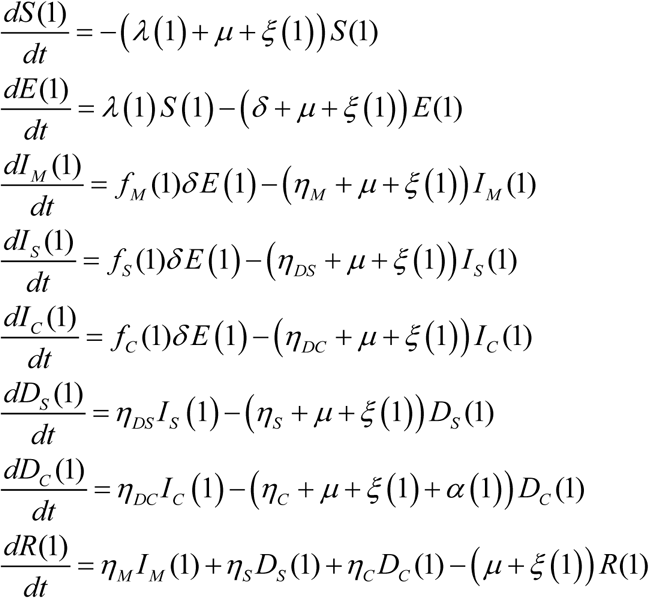

For subsequent age groups, the following set of equations was used:

<u>Populations aged 10+ years:</u>

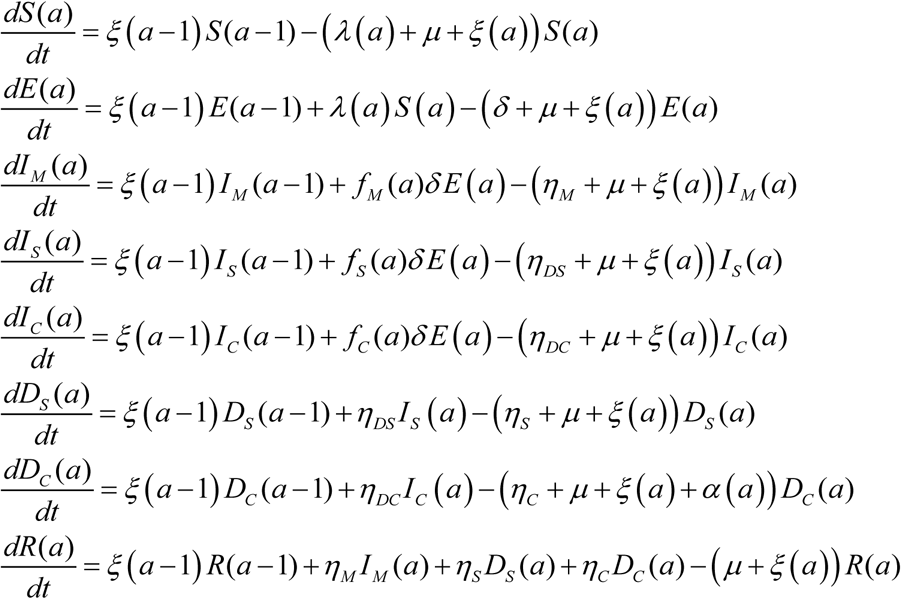

The definitions of population variables and symbols used in the equations are listed in Table S1.

**Table S1.**
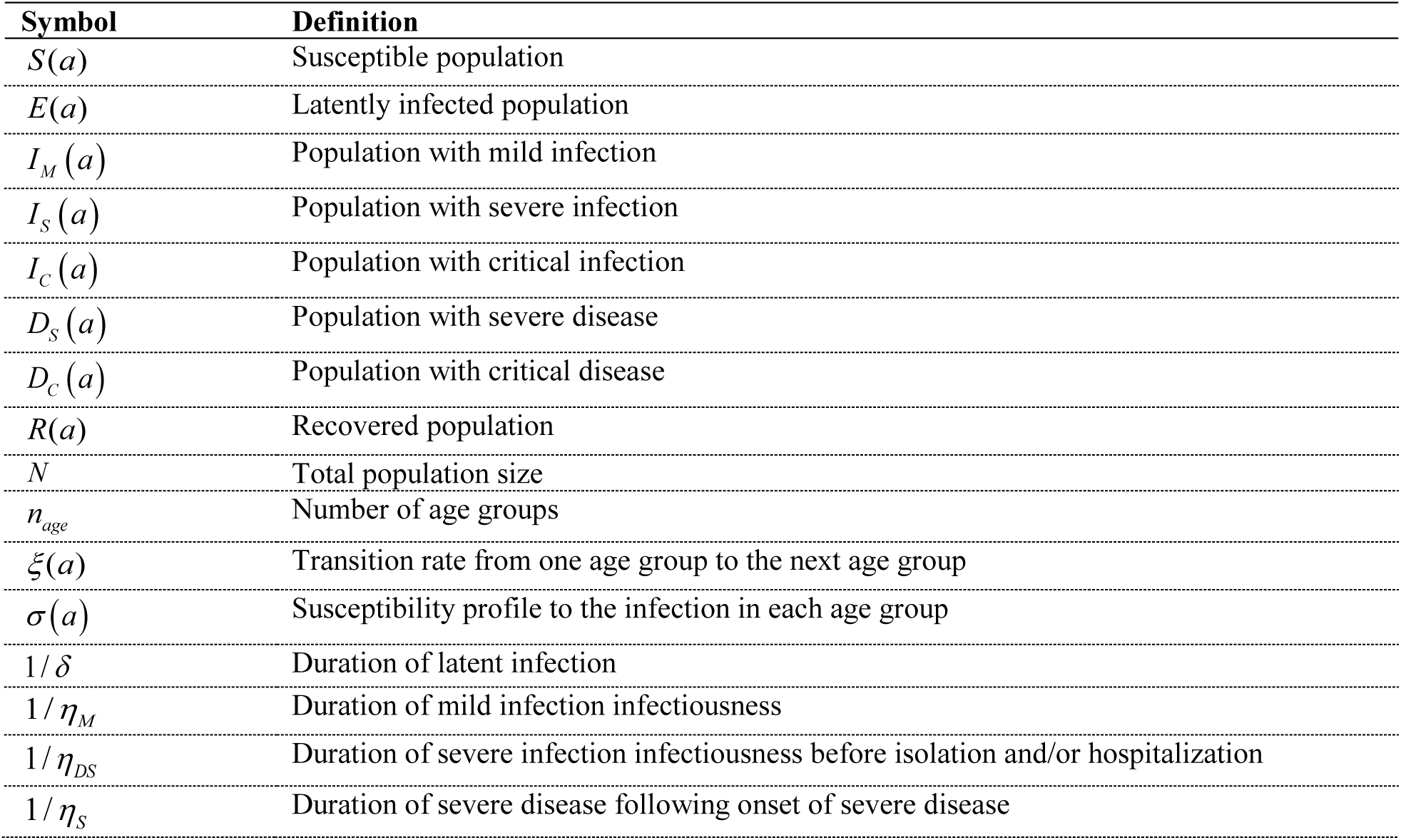
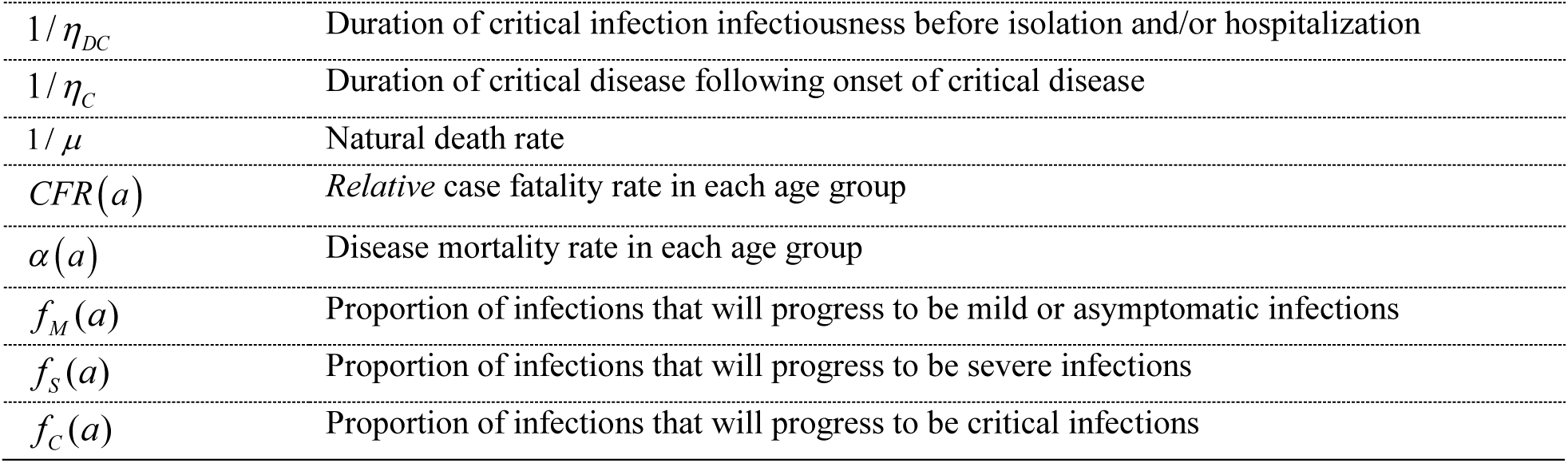
Definitions of population variables and symbols used in the model.

| Symbol | Definition |
| --- | --- |
| $S(a)$ | Susceptible population |
| $E(a)$ | Latently infected population |
| $I_M(a)$ | Population with mild infection |
| $I_S(a)$ | Population with severe infection |
| $I_C(a)$ | Population with critical infection |
| $D_S(a)$ | Population with severe disease |
| $D_C(a)$ | Population with critical disease |
| $R(a)$ | Recovered population |
| $N$ | Total population size |
| $n_{age}$ | Number of age groups |
| $\xi(a)$ | Transition rate from one age group to the next age group |
| $\sigma(a)$ | Susceptibility profile to the infection in each age group |
| $1/\delta$ | Duration of latent infection |
| $1/\eta_M$ | Duration of mild infection infectiousness |
| $1/\eta_{DS}$ | Duration of severe infection infectiousness before isolation and/or hospitalization |
| $1/\eta_S$ | Duration of severe disease following onset of severe disease |
| $1/\eta_{DC}$ | Duration of critical infection infectiousness before isolation and/or hospitalization |
| $1/\eta_C$ | Duration of critical disease following onset of critical disease |
| $1/\mu$ | Natural death rate |
| $CFR(a)$ | *Relative* case fatality rate in each age group |
| $\alpha(a)$ | Disease mortality rate in each age group |
| $f_M(a)$ | Proportion of infections that will progress to be mild or asymptomatic infections |
| $f_S(a)$ | Proportion of infections that will progress to be severe infections |
| $f_C(a)$ | Proportion of infections that will progress to be critical infections |

The force of infection (hazard rate of infection) experienced by each susceptible population *S* (*a*) is given by

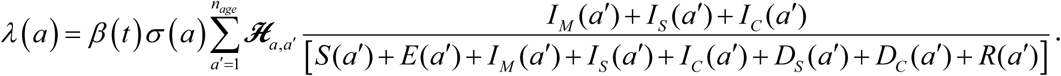

Here, *β* (*t*) is the rate of infectious contacts parameterized to capture the effect of the public health interventions implemented in China through a Woods-Saxon function [3-6]. This function is mathematically designed to describe and characterize transitions in terms of their scale or strength, smoothness or abruptness, thickness (duration), and the turning point [3, 4]:

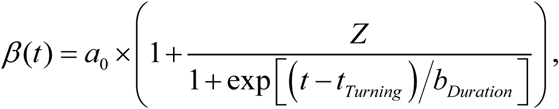

where *a*_0_, *Z, t*_*Turning*_, *b*_*Duration*_ are the fitting parameters used to describe the reduction in the rate of infectious contacts.

The mixing among the different age groups is dictated by the mixing matrix **ℋ**_*a, a*′_ This matrix provides the probability that an individual in the *a* age group will mix with an individual in the *a*′ age group. The mixing matrix is given by

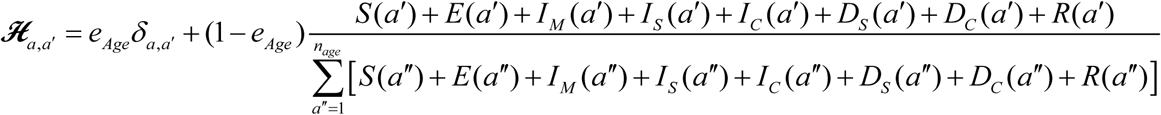

Here, *δ*_*a,a*′_; is the identity matrix. *e*_*Age*_ ∈[0,1] measures the degree of assortativeness in the mixing. At the extreme *e*_*Age*_ = 0, the mixing is fully proportional. Meanwhile, at the other extreme, *e*_*Age*_ = 1, the mixing is fully assortative, that is individuals mix only with members in their own age group.

The disease mortality rate *α* (*a*) was parametrized through an overall fitting factor multiplied by the observed crude age-stratified case fatality rate.

## 2. Parameter values

The input parameters of the model were chosen based on current empirical data for COVID-19 natural history and epidemiology. The parameter values are listed in Table S2.

**Table S2.** Model assumptions in terms of parameter values.

| Parameter | Symbol | Value | Justification |
| --- | --- | --- | --- |
| Duration of latent infection | $1/\delta$ | 3.69 days | Based on existing estimate [7] and based on a median incubation period of 5.1 days [8] adjusted by observed viral load among infected persons [9] and reported transmission before onset of symptoms [10] |
| Duration of infectiousness | $1/\eta_M$ ;<br>$1/\eta_{DS}$ ;<br>$1/\eta_{DC}$ | 3.48 days | Based on existing estimate [7] and based on observed time to recovery among persons with mild infection [7, 11] and observed viral load in infected persons [9, 10] |
| Duration of severe disease following onset of severe disease | $1/\eta_S$ | 28 days | Observed duration from onset of severe disease to recovery [11] |
| Duration of hospitalization for critical infection | $1/\eta_C$ | 28 days | Observed duration from onset of critical disease to recovery [11] |
| Life expectancy in China | $1/\mu$ | 77.47 years | United Nations World Population Prospects database [1] |
| Crude case fatality rate in each age group | $CFR(a)$ | | Observed crude case fatality rate based on China data as of February 11, 2020 [2, 12] |
| Age 0-9 years |  | 0% |  |
| Age 10-39 years |  | 0.2% |  |
| Age 40-49 years |  | 0.4% |  |
| Age 50-59 years |  | 1.3% |  |
| Age 60-69 years |  | 3.6% |  |
| Age 70-79 years |  | 8.0% |  |
| Age 80+ years |  | 21.9% |  |
| Proportion of infections that will progress to be mild or asymptomatic infections | $f_M(a)$ | | Observed proportion of infections that eventually develop mild or asymptomatic in China [11, 13, 14] |
| Age 0-9 years |  | 88.9% |  |
| Age 10-49 years |  | 88.0% |  |
| Age 50-59 years |  | 82.5% |  |
| Age 60+ years |  | 71.2% |  |
| Proportion of infections that will progress to be severe infections | $f_S(a)$ | | Observed proportion of infections that eventually develop severe disease in China [11, 13, 14] |
| Age 0-9 years |  | 11.1% |  |
| Age 10-49 years |  | 9.9% |  |
| Age 50-59 years |  | 10.3% |  |
| Age 60+ years |  | 7.8% |  |
| Proportion of infections that will progress to be critical infections | $f_C(a)$ | | Observed proportion of infections that eventually develop critical disease in China [11, 13, 14] |
| Age 0-9 years |  | 0.0% |  |
| Age 10-49 years |  | 2.2% |  |
| Age 50-59 years |  | 7.2% |  |
| Age 60+ years |  | 20.9% |  |

## 3. Adjustment for the outlying values for the number of cases and deaths reported on February 13, 2020

There was a change in coronavirus case definition in China on February 13, 2020, to include, in addition to the laboratory-confirmed cases, those who are clinically-diagnosed [15, 16]. This led to a very sharp one-day increase in the reported numbers of infections and deaths, respectively [17]. These “outlying observations” included the “excess” cases not accounted for in previous days.

To correct for this data artifact, we distributed the number of cases reported on February 13, 2020 over a time duration including this date and previous dates. This was done through the condition:

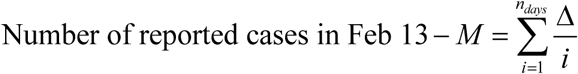

Here, Δ/*i* is the increment added to the reported number of cases in each *i* -day starting with February 13 and moving backward, *M* + Δ is the adjusted number of cases in February 13, *M* and *n*_*days*_ are fitting parameters, and Δ calculated using the above constraint. Model fitting (and uncertainty analysis) yielded the following estimates: *M* = 1,187 (95% uncertainty interval (UI): 970-1,453) and *n*_*days*_ = 25 (95% UI: 22-29) days.

Similarly, we distributed the number of deaths reported on February 13, 2020 over a time duration including this date and previous dates. This was done through the condition:

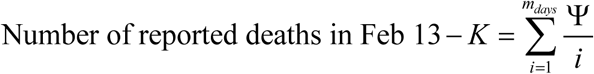

Here, Ψ/*i* is the increment added to the reported number of cases in each *i* -day starting with February 13 and moving backward, *K* + Ψ is the adjusted number of cases in February 13, *K* and *m*_*days*_ are fitting parameters, and Ψ calculated using the above constraint. Model fitting (and uncertainty analysis) yielded the following estimates: *K* = 109 (95% UI: 104-114) and *m*_*days*_ = 16 (95% UI: 13-20) days.

**Figure S2.**
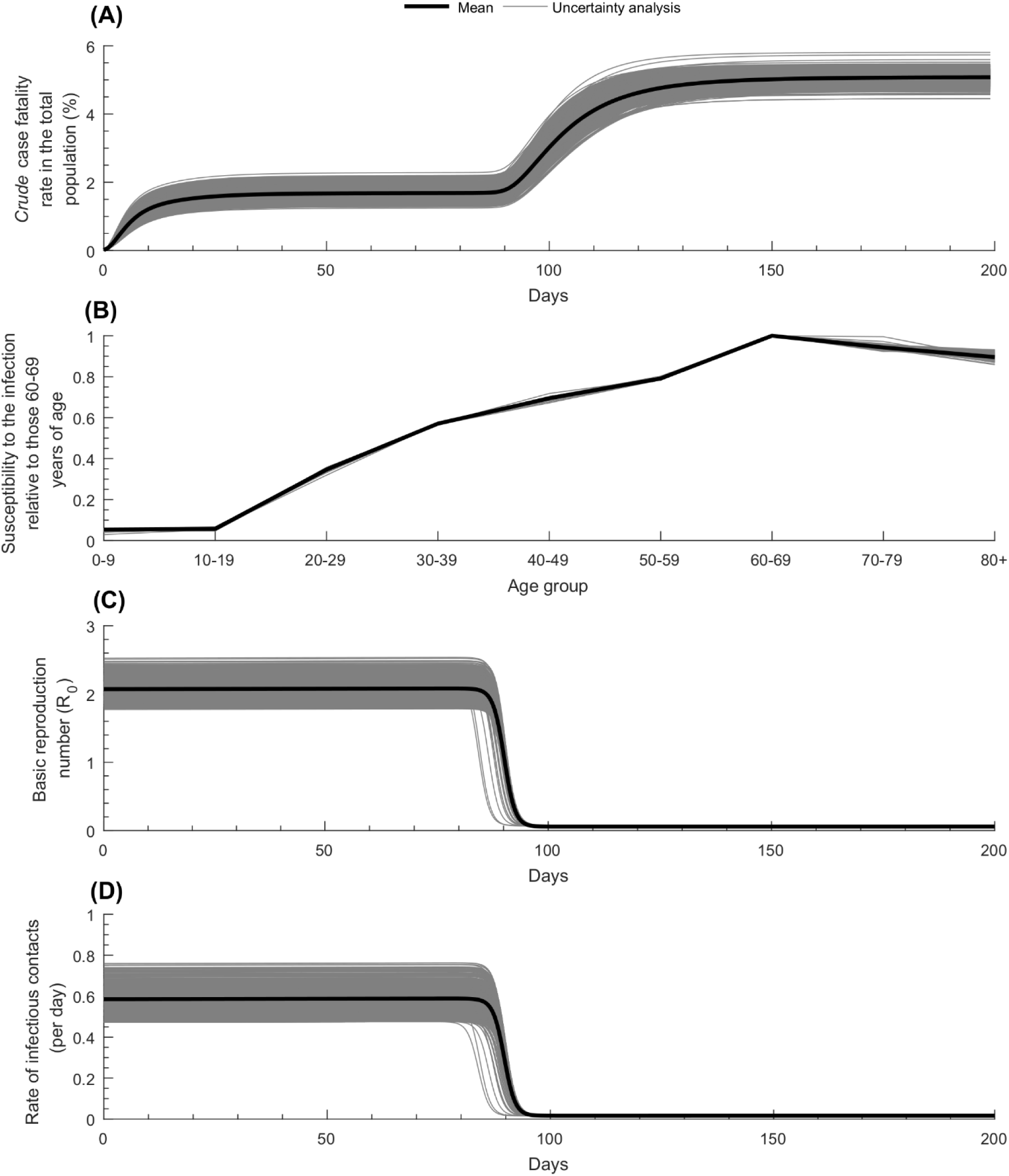
Uncertainty analysis estimating the range of uncertainty around key model predictions. Results of 500 uncertainty runs showing (A) the time-dependent crude case fatality rate in the total population, (B) the age-dependence of the relative susceptibility to the infection, (C) the time-dependent basic reproduction number, and (D) the time-dependent rate of infectious contacts.

**Figure S3.**
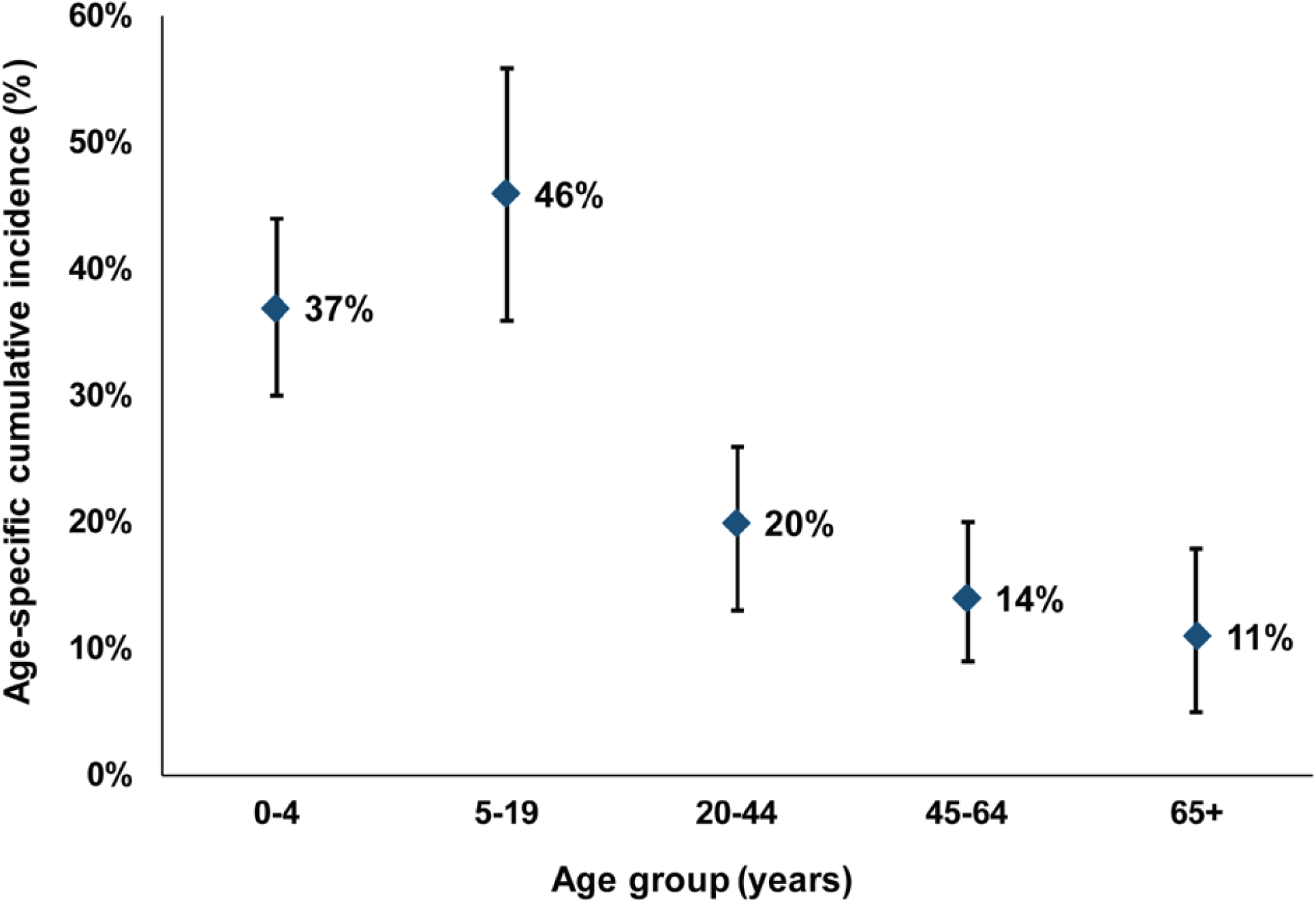
Age-specific cumulative incidence of the 2009 influenza A (H1N1) pandemic (H1N1pdm) virus [18].

